# Coping with Cannabis During Pregnancy: Trajectories of Depression, Stress, and Cannabis Use across the Prenatal Period

**DOI:** 10.1101/2024.04.09.24305545

**Authors:** Anna Constantino-Pettit, Rebecca Tillman, Jillian Wilson, Nicole Lashley-Simms, Naazanene Vatan, Azaria Atkinson, Shelby Leverett, Shannon Lenze, Christopher Smyser, Ryan Bogdan, Cynthia Rogers, Arpana Agrawal

## Abstract

**Objective:** We examined trajectories of stress, depression, and cannabis use across the prenatal period. We also investigated whether individuals who reported using cannabis to alleviate stress and depressive symptoms experienced symptom relief across the prenatal period.

**Methods:** Pregnant individuals (n=436) with a history of lifetime cannabis use were recruited and identified either as prenatal cannabis users (PCU; continued cannabis use following knowledge of pregnancy in the first trimester) or non-prenatal cannabis users (NPCU; no cannabis use following knowledge of pregnancy in the first trimester). PCU individuals additionally reported on reasons for continued cannabis use during pregnancy. We employed longitudinal multivariate modeling to examine trajectories of depression (maternal self-report; Edinburgh Postnatal Depression Scale), stress (maternal self-report; Perceived Stress Scale), and cannabis use (maternal self-report and urinalysis) over time, stratified by motives for use during the perinatal period.

**Results:** Stress, depression, and cannabis use decreased from the first to the third trimester (slope *B=* -0.21, -0.35, and -0.31, respectively). While cannabis use and depression at the first trimester were correlated with one another, they did not affect the other’s rate of change. Cannabis use similarly did not affect the rate of change in stress. Finally, while individuals using cannabis to cope with mental health symptoms experienced a decrease in overall depression symptoms, this slope was roughly equivalent to individuals who were not using cannabis prenatally (slope *B=* - 0.43 and -0.51, respectively).

**Conclusion:** Our sample experienced a collective decrease in depression, stress, and cannabis use prenatally. This decline in cannabis use – even among those who reported using to cope with mental health conditions – was not attributable to the decrease in depressive symptoms. This study lends valuable insight into the reasons why individuals continue to use cannabis during pregnancy, which providers can use to help tailor recommendations for other sources of coping and support to childbearing individuals.

## Introduction

The past ten years have seen a marked increase in cannabis use, particularly in states where legislation has passed to make both medical and recreational marijuana legal (Hammond et al. 2020). Given the recent legislative changes, along with proximal environmental stressors including the COVID-19 pandemic, it is important to understand whether cannabis use behavior has changed among vulnerable populations, particularly pregnant individuals. Much of the available data on prenatal cannabis use in the US has been cross-sectional, and national estimates have reported rates of perinatal cannabis use at around 5% (Alshaarawy & Vanderziel, 2022). However, recent research has taken advantage of national-level data registries including the Pregnancy Risk Assessment Monitoring System (PRAMS) or the National Survey on Drug Use and Health (NSDUH) to examine trends of prenatal cannabis use. Data from PRAMS has shown that pre-conception and postpartum, but not prenatal, cannabis use increased among individuals in states with legislation of recreational cannabis use (Skelton et al. 2021). Data from NSDUH, which did not differentiate between individuals living in states with versus without cannabis legalization, has shown an overall increase for prenatal cannabis use from around 2.85% in 2002 to nearly 5% in 2016 (Agrawal et al., 2018). Further, the relative amount of tetrahydrocannabinol (THC), which is the main psychoactive component of cannabis, has increased substantially in recent years (Volkow et al., 2019). Other studies have reported on local increases in prenatal cannabis use; for instance, Young-Wolff et al. found a 25% increase in prenatal cannabis use during the pandemic among California residents (2021). Based on the available data, the prevalence of use at some point during the perinatal period is estimated to be around 10-20% in high-income countries such as the United States (Singh et al. 2020).

While there is evidence, therefore, of an increase in cannabis use during the prenatal period, we have little insight into why pregnant individuals continue to use cannabis during pregnancy. Previous studies have reported social and behavioral characteristics of pregnant individuals who are more likely to use cannabis, albeit without the motive. Characteristics include having a partner who used cannabis during their pregnancy, not being in a relationship, having a trauma history, and concurrent use of tobacco, alcohol, or illicit drugs (El Marroun et al., 2008; Ko et al., 2015). One theory is that perinatal individuals are using cannabis to cope with the various symptoms that frequently accompany pregnancy, including mental health symptoms. Mental health trajectories, similar to cannabis use trajectories, have appeared to increase at a population level during the prenatal period over the past ten years. Prenatal depression, which continues to be a leading complication of pregnancy, has been increasing generationally, with global estimates hovering around 11% (Pearson et al. 2018; Woody et al. 2017). This increase in prenatal depression is likely multifactorial; the American College of Obstetricians and Gynecologists (ACOG) recommended universal screening for perinatal depression in 2016, which has since led to a decrease in stigma associated with the condition (Fedock et al., 2018). Increased rates of prenatal depression have also been associated with more recent environmental stressors such as the COVID-19 pandemic (Mateus et al., 2022). Similarly, prenatal stress has steadily risen in prevalence estimates generationally (Rubertsson et al. 2014). Prenatal stress regularly co-occurs with prenatal depression, and is believed to affect about 20% of childbearing individuals (Dennis et al., 2018). Similar to prenatal depression, prenatal stress affects both the pregnant individual and their infant: prenatal stress has been associated with poverty and interpersonal violence (Field 2017), as well as preterm birth and low birth weight (Ding et al., 2014).

Some extant studies have attempted to classify reasons for cannabis use during pregnancy, but different methodologies and operationalization around prenatal cannabis use has made it difficult to draw reliable conclusions about prenatal individuals’ motives for use. For instance, one qualitative study examined reasons for cannabis use among individuals who had been pregnant in the past year and found that individuals who used during pregnancy did so primarily to manage physical symptoms such as nausea and vomiting (Vanstone et al., 2021). Another recent qualitative study found that pregnant individuals primarily cited physical health concerns, including pain, as reasons for continued cannabis use during pregnancy. In fact, this study found that multiple individuals expressed concerns about potential risk to the fetus from traditional prescribed or over-the-counter pain medications, and reasoned that cannabis was a safer choice (Kiel et al., 2023). Besse et al. found a similar rationale for continued use among a clinically-ascertained sample of prenatal individuals, although the physical symptoms frequently cited among their prenatal cannabis users included sleep issues (2023). This sample also reported using cannabis to manage relaxation, anxiety, and depression. It is unclear whether individuals are weighing the risks and benefits of using cannabis versus psychotropic medications – similar to individuals who have stated that they chose to use cannabis for pain rather than other medications, which they perceived as more harmful to their unborn baby – because few studies have published data on the overlap between cannabis use and psychotropic medication utilization in this population. One cross-sectional study found that the majority of individuals who cited using cannabis during pregnancy for mental health reasons were not taking psychotropic medication (Regalado et al., 2023).

Importantly, no study to date has attempted to examine the efficacy of cannabis use to decrease mental health symptoms, among those who cite using cannabis prenatally primarily for mental health purposes. To that end, we examined trajectories of stress, depression, and cannabis use across the prenatal period. We were interested in answering the following questions; first, what are the patterns of change from the first to the third trimester in stress, depression and cannabis use? And second, among those who are continuing to use to mitigate the effects of anxiety and depression, is there a perceptible benefit from cannabis use on mental health symptoms?

## Methods

### Sample

Data for the current study was taken from the Cannabis Use During Early Life and Development (CUDDEL) study. This sample, recruited from a Midwest regional medical center obstetrics clinic, was recruited based on a lifetime history of cannabis use prior to conception.

Once individuals learned of their pregnancy during the first trimester, they self-reported either cannabis cessation (controls / NPCU group; n=152) or continued cannabis use (cases / PCU group; n=284). Importantly, to be eligible for the study, individuals could not have concurrent tobacco use, illicit drug use, or heavy alcohol use. Self-reported cannabis use (or non-use) was substantiated with the addition of a urine drug screen (UDS) at each trimester. For a complete description of CUDDEL recruitment strategies and protocols, see Bogdan et al. 2024. This study was approved by the Washington University School of Medicine institutional review board (ID 20180001).

### Measures

#### Prenatal Depression and Stress

Prenatal depression was measured using the Edinburgh Postnatal Depression Scale (EPDS; Cox et al. 1987) while prenatal stress was ascertained using the Perceived Stress Scale (PSS; Cohen et al. 1994). Both measures were completed once during each trimester. .

#### Prenatal Cannabis Use

Prenatal cannabis use (PCU) was ascertained both via participant self-report and verified with a UDS at each trimester. In the baseline survey, which participants completed during their first trimester of pregnancy, they were asked the following question: “During this pregnancy, how often did you use marijuana after you knew that you were pregnant?”. Responses included “Never” (0); “Monthly or less” (1); “2-4 times a month” (2); “2- 3 times a week” (3); or “4 or more times a week” (4). Participants were asked the same question “Since your last study visit…” for trimesters two and three. Cases (PCU) were coded as any positive responses to the self-report questions, whereas controls (nPCU) were coded as any who responded “Never” at each of the three trimesters. Participants additionally completed a UDS at each trimester. UDS were completed using urine drug testing cups and strips (Test Country, 12 panel cup; Confirm Biosciences 12 panel cup/DrugConfirm; 12panelnow 10 panel cup; Premier Brands cotinine strip) and tested for the following substances: cannabis, nicotine (cotinine), amphetamines, benzodiazepines, buprenorphine, cocaine, ecstasy, methamphetamine, methadone, morphine, and oxycodone. In any cases where there was a discrepancy between a positive UDS and self-report, participants were recorded as a case (PCU).

#### Coping and Other Reasons for Use

During the first trimester, participants who responded positively to using cannabis following knowledge of the pregnancy were asked “Different women have different reasons for using marijuana during pregnancy. Please mark all that apply to <u>you</u>”. Responses included: “To deal with nausea/vomiting”; “To deal with anxiety or stress”; “To help with hunger or loss of appetite”; “To help you sleep”; “To treat mental health problems (e.g. depression, posttraumatic stress disorder)”; “For enjoyment and relaxation”; “To have energy, get more things done during the day”; “To help with pain”; “It was uncomfortable to stop using”; “Other”. Any individuals who marked either “To deal with anxiety or stress” or “To treat mental health problems (e.g. depression, posttraumatic stress disorder)” were classified as COPE participants – participants who were using to cope with mental health symptoms.

#### Income-to-Needs

Income-to-needs ratios were derived by dividing total household income by the federal guideline for poverty, given the size of the family, in the year the data was collected. *History of Mental Health*. Mental health history was ascertained from the participants’ electronic health records; any mental health diagnoses that were present and active in the participant’s chart at the time of their intake were recorded and transformed into a binary variable (1= mental health history; 0 = none).

#### Psychotropic Medication

Psychotropic medication was ascertained from a review of the participants’ electronic health records during their first trimester. Psychotropic medication was coded as a binary variable (1= psychotropic medication use during the first trimester; 0=none).

### Data analysis

We estimated mean EPDS and PSS across trimesters and as a function of prenatal cannabis use (PCU), which was defined as a positive self-report or urine drug screen for cannabis at any trimester, and in those self-reporting use at their 1st trimester visit, as a function of whether they were using cannabis to cope with stress or anxiety or to deal with a psychiatric condition (COPE). Similarly, mean cannabis use at each trimester was calculated. Pairwise comparisons of means for PCU versus nPCU were conducted using t-tests in SAS. All subsequent growth models were fitted to data in Mplus v8. First, to estimate change in EPDS, PSS and PCU, linear growth curve models were fitted, individually, to EPDS and PSS data across the 3 trimesters. For each analysis, we included a second covariate adjusted model, where the intercept and slope were regressed on age at screening, income-to-needs ratio, a history of mental health problems, or evidence of psychotropic medication usage. Next, a parallel process growth model was fitted to (a)EPDS and PCU, and (b) PSS and PCU, to jointly estimate change in prenatal cannabis use alongside EPDS or PSS. Finally, to model changes in EPDS and PSS across participants without a history of PCU and within those with PCU, by COPE, a 3-group linear growth model was fitted to data on EPDS and PSS. Intercepts and slopes were freely estimated in each group (i.e., nPCU, PCU-COPE PCU+COPE) and any non-significant estimates were constrained to zero for parsimony, followed by sequential equality constraints on the intercepts and slopes. Overall model-fit was evaluated using the Comparative fit index (CFI) > 0.95 and root mean square error of approximation (RMSEA) < 0.06, which are generally considered an indication of good fit (Bollen & Curran, 2006). Changes in model fit were calculated using the difference in chi-square for given degrees of freedom.

## Results

Sample demographics can be found in Table 1. The majority of participants self-identified as Black with an average age of 26 and average INR of 1.2. There were some modest but significant differences in demographic characteristics between PCU and nPCU participants. PCU participants were, on average, about a year younger than nPCU (PCU 25.8, nPCU 27.0; *t*=2.6, p=.009). Both were affected by poverty, although the PCU group was at the poverty line (PCU 1.0, nPCU 1.6; *t*=5.3, p<.001). While 86% of our sample was Black, there was a greater proportion of Black participants in the PCU compared to the nPCU group (PCU 89%, nPCU 80%; *x*^2^=7.7, p=.006). There were no significant differences in PCU compared to the nPCU group in depression or stress by trimester. The PCU group reported higher average stress at trimesters 2 and 3, but this only approached significance (p=.06 and p=.08, respectively).

**Table 1.** Sample Demographics.

|  | **Total**<br>**(n = 464)** | **No PCU**<br>**(n = 161)** | **PCU**<br>**(n = 303)** | **No PCU vs.**<br>**PCU** |
| --- | --- | --- | --- | --- |
| **Variable** | **Mean (SD)** | **Mean (SD)** | **Mean (SD)** | **p** |
| Trimester 1 EPDS | 7.8 (5.8) | 8.1 (6.0) | 7.6 (5.7) | 0.40 |
| Trimester 2 EPDS | 6.7 (5.7) | 6.4 (5.9) | 6.8 (5.5) | 0.53 |
| Trimester 3 EPDS | 6.1 (5.4) | 6.0 (5.1) | 6.3 (5.5) | 0.60 |
| Trimester 1 PSS | 16.9 (7.7) | 16.3 (7.9) | 17.0 (7.7) | 0.39 |
| Trimester 2 PSS | 15.7 (7.3) | 14.7(7.8) | 16.3 (7.0) | 0.06 |
| Trimester 3 PSS | 15.1 (7.0) | 14.2 (7.1) | 15.7 (6.9) | 0.08 |
| Age | 26.3 (4.9) | 27.0 (5.2) | 25.8 (4.7) | **0.00** |
| INR | 1.2 (1.0) | 1.6 (1.5) | 1.0 (0.5) | **0.00** |
|  | **% (n)** | **% (n)** | **% (n)** | **p** |
| Race |  |  |  |  |
| Black | 86.2 (400) | 80.1 (129) | 89.4 (271) | **0.00** |
| Other | 13.8 (64) | 19.9 (32) | 10.6 (32) |  |

Reasons for cannabis use can be found in Figure 1. Participants primarily reported using cannabis to manage nausea and vomiting (60%) associated with the pregnancy. The second-highest endorsed reason for use was, relatedly, to manage hunger and appetite (53%). Participants rated stress (46%) higher than mental health needs (24%) as reasons for continued use, but also reported using for both sleep (41%) and pain (28%), which frequently co-occur with depressive disorders.

**Figure 1.**
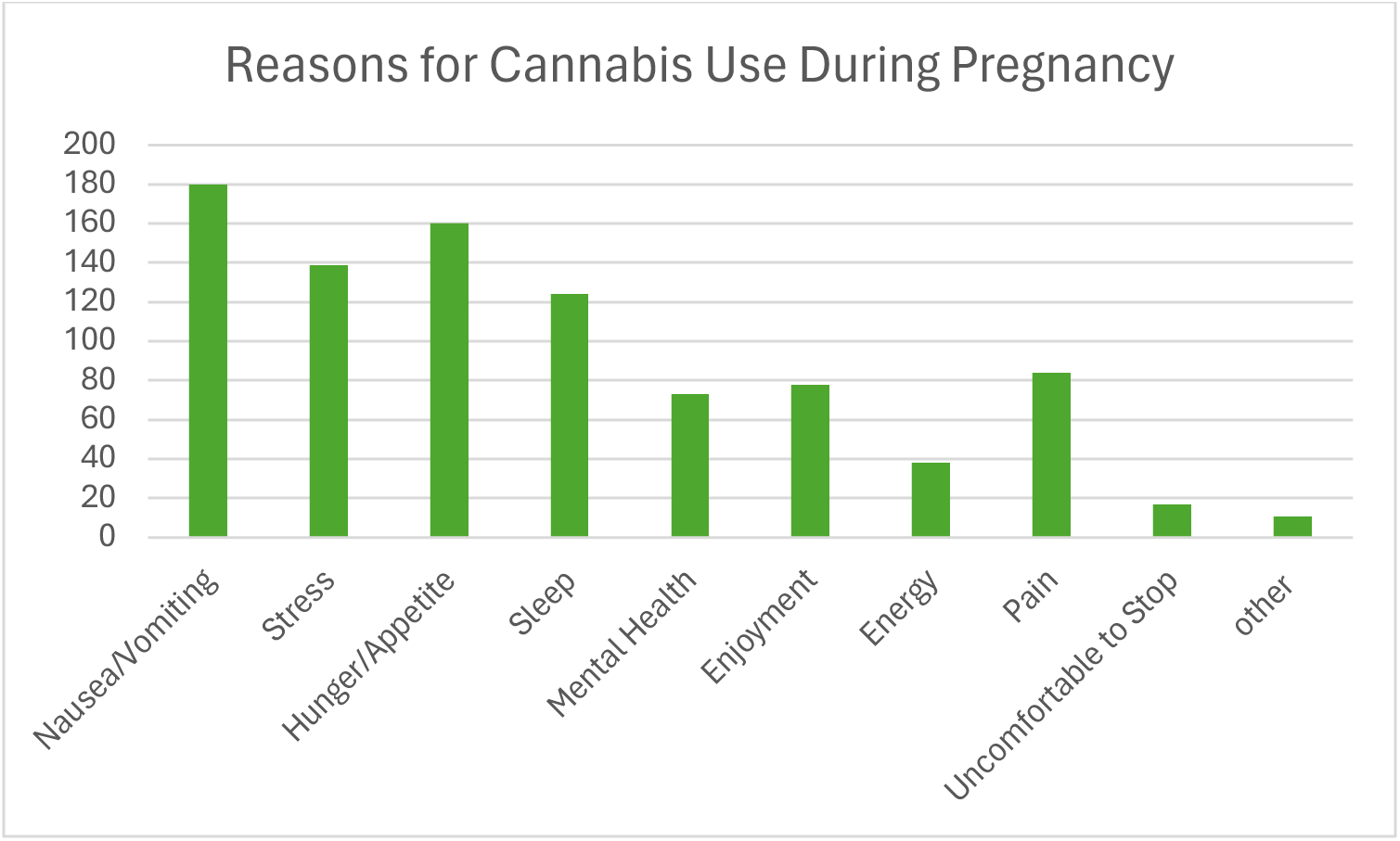

**Figure 2.**
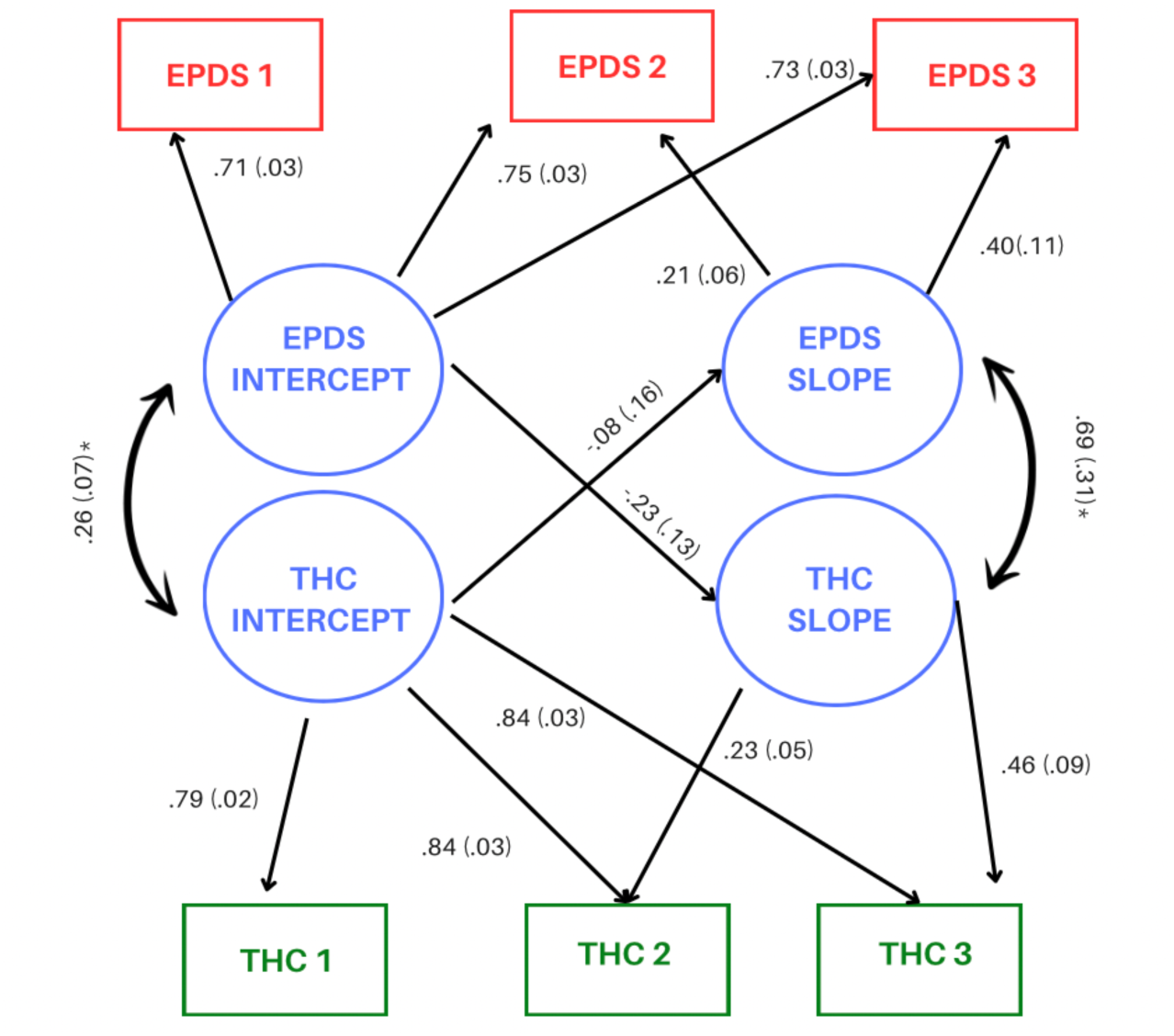
Dual Trajectory Model: EPDS and Cannabis Use (n=504)

Baseline and Covariate-Adjusted Models: Table 2 presents the results for both the baseline and covariate-adjusted trajectories for cannabis use, stress, and depression across the prenatal period. A baseline model describing the trajectory of cannabis use across pregnancy fit the data well (CFI 1.00, *x*^2^= 0.11, df = 1). Mean cannabis use decreased across trimesters, as did depression (CFI = 0.998, *x*^2^=304.14, df = 3) and stress (CFI 1.00, *x*^2^=277.52, df = 1). Slopes were correlated with intercepts for depression, stress, and cannabis use (set at trimester 1; Table 2). Regressing the slope and intercept on covariates did not alter findings. For PCU, age (*B*=-0.11., p=0.04) and INR (*B*=-0.14, p=0.01) were negatively associated with T1 use. For Prenatal Depression, baseline endorsement of mental health problems (*B*=0.19, p <0.0001) and a mental health history (*B*=0.34, p = 0.00) were associated with higher T1 EPDS scores. Mental health history alone was significantly negatively associated with the decreasing rate of change in depression scores (*B*=- 0.20; p = 0.03). Age was also significantly negatively associated with decreased depression scores at T1 (*B*=-0.11, p =0.04). For Stress, mental health history (*B*=0.23, p=0.00) and use of psychotropic medication (*B*=0.19, p=0.003) were positively associated with higher symptoms at T1, but were not associated with the rate of change. Similar to depression, age was also negatively associated with a decrease in stress symptoms at T1 (*B*= -0.15, p = 0.002).

**Table 2.** **Baseline Model:** Standardized Estimates and 95% Confidence Intervals For Cannabis, Stress, and Depression Trajectories; **Covariate-Adjusted Model**: Standardized Estimates for Covariates in the Models

| Baseline Model | Prenatal Cannabis Use (n=504) |  |  | Prenatal Stress (n=417) |  |  | Prenatal Depression (n=478) |  |  |
| --- | --- | --- | --- | --- | --- | --- | --- | --- | --- |
|  | Intercept | Slope | Slope-Intercept Correlation | Intercept | Slope | Slope-Intercept Correlation | Intercept | Slope | Slope-Intercept Correlation |
|  | Std. Est.<br>[95% CI] | Std. Est.<br>[95% CI] | Std. Est.<br>[95% CI] | Std. Est.<br>[95% CI] | Std. Est.<br>[95% CI] | Std. Est.<br>[95% CI] | Std. Est.<br>[95% CI] | Std. Est.<br>[95% CI] | Std. Est.<br>[95% CI] |
| No covariates | 0.90<br>[0.79, 1.01] | -0.31<br>[-.44, -.19] | -0.35<br>[-.47, -.23] | 2.59<br>[2.31, 2.87] | -0.29<br>[-.43, -.15] | -0.49<br>[-.62, -.36] | 1.58<br>[1.40, 1.77] | -0.35<br>[-.49, -.21] | -0.45<br>[-.58, -.31] |
| Covariate-Adjusted Model | Prenatal Cannabis Use (n=405) |  |  | Prenatal Stress (n=372) |  |  | Prenatal Depression (n=397) |  |  |
|  | Intercept | Slope | Slope-Intercept Correlation | Intercept | Slope | Slope-Intercept Correlation | Intercept | Slope | Slope-Intercept Correlation |
|  | Std. Est. | Std. Est. | Std. Est. | Std. Est. | Std. Est. | Std. Est. | Std. Est. | Std. Est. | Std. Est. |
|  |  |  | -0.35*** |  |  | -0.51*** |  |  | -0.39*** |
| Age | -0.11* | 0.05 |  | -0.15** | 0.07 |  | -0.11* | 0.01 |  |
| INR | -0.14* | 0.07 |  | -0.02 | -0.08 |  | 0.03 | -0.03 |  |
| Mental Health History | 0.01 | 0.07 |  | 0.23*** | 0.00 |  | 0.34*** | -0.20* |  |
| Psychotropic Medication | 0.05 | -0.04 |  | 0.19** | -0.11 |  | 0.19** | -0.10 |  |

### Dual-Trajectory Models

Table 3 presents the results for the dual-trajectory model of cannabis use and stress (CFI 0.97, *x*^2^ = 29.8, df = 9), while Figure 2 presents the results for the dual-trajectory model of cannabis use and depression (CFI 0.99, *x*^2^ = 22.0, df = 9). Intercepts, but not slopes, were correlated across prenatal cannabis use and stress. Covariate effects (*x*^2=^33.13, df= 5, CFI= 0.98) on intercepts were identical to those noted for prenatal depression but, in contrast, baseline mental health history did not modify either slope. T1 cannabis use and depressive symptoms were correlated as was the rate of change (i.e., decline, negative slopes) of both behaviors. However, there was no evidence for cross-variable effects such that T1 cannabis use did not influence the rate of change in depressive symptoms during pregnancy nor did T1 depressive symptoms scores influence rate of change in cannabis use from T1 to T3. In the covariate-adjusted model, which did not appreciably improve model fit (*x*^2^=25.31, df= 15, CFI = 0.99), age and baseline mental health history were significantly associated with baseline depressive symptoms (Age: *B* =-0.13, SE=0.06, p=0.03; Mental Health: *B* =0.46, SE=0.05, p <0.0001), while age and INR were significantly associated with baseline cannabis use (Age: *B* =- 0.12, SE=0.06, p=0.03; INR: *B* = -0.15, SE=0.06, p=0.01). In addition, T1 mental health was negatively associated with the slope for depression (*B* =-0.36, SE=0.12, p=0.002) and nominally, positively with the prenatal cannabis slope (*B* =0.22, SE=0.12, p=0.05).

**Table 3.** Dual Trajectory Model: PSS and Cannabis Use (n=504)

|  | Std. Est. [95% CI] |
| --- | --- |
| Stress | 3.11 [2.82, 3.40] |
| Trim 1 – Intercept | 0.70 [0.66,0.75] |
| Trim 2 – Intercept | 0.77 [0.71,0.82] |
| Trim 3 – Intercept | 0.75 [0.70,0.81] |
| Trim 1 – Slope | 0.00 [0.00,0.00] |
| Trim 2 – Slope | 0.14 [-0.02,0.29] |
| Trim 3 – Slope | 0.27 [-0.03,0.57] |
| Cannabis | 1.00 [0.89, 1.12] |
| Trim 1 – Intercept | 0.78 [0.74,0.82] |
| Trim 2 – Intercept | 0.84 [0.80,0.88] |
| Trim 3 – Intercept | 0.82 [0.78,0.87] |
| Trim 1 – Slope | 0.00 [0.00,0.00] |
| Trim 2 – Slope | 0.22 [0.14,0.30] |
| Trim 3 – Slope | 0.44 [0.28,0.59] |
| Stress slope on Cannabis intercept | -0.01 [-0.44, 0.41] |
| Cannabis slope on Stress intercept | -0.14 [-0.37, 0.09] |
| Cannabis intercept with Stress intercept | 0.26 [0.13, 0.40] |
| Cannabis slope with Stress slope | -0.24 [-0.98, 0.50] |

#### Reasons for Use (COPE) Models

Table 5 presents the dual trajectories of cannabis use and depression symptoms among participants who cited using cannabis to alleviate mental health symptoms (CFI 0.97, x^2^=12.75, df=6). As expected, T1 depression scores (intercepts) were statistically different across the 3 groups. There was no statistical change in depression scores in the PCU-COPE group; we therefore constrained that slope to 0 in the final models. The rate of change in depression scores from T1 to T3 was roughly equivalent across those who did not use cannabis during pregnancy (nPCU; *B* = -0.43) and those who used it and reported anxiety/stress/psychiatric symptoms as their motive for use (PCU+COPE; *B* = -0.51). Table 6 presents the dual trajectories of cannabis use and stress symptoms among participants who cited using cannabis to alleviate mental health symptoms (CFI=0.99, x^2^=7.42, df=6). Similarly, for prenatal stress, T1 scores were statistically different across groups while change in stress scores across time did not differ substantially (nPCU *B* = -0.43; PCU+COPE *B* = -0.32). Thus, for both prenatal depression and stress, while T1 scores differed at baseline (intercepts), there was no change in these symptoms across pregnancy for those using cannabis but not reporting COPE (PCU-COPE). Furthermore, the rate of change in prenatal depression and stress was equivalent in those who did not use cannabis during pregnancy (nPCU) and those who used it to cope (PCU+COPE).

**Table 5.** COPE Model 1: Does Depression Decrease Among Moms Using Cannabis to Cope with Mental Health Difficulties?

|  | **G1: No Cannabis Use**<br>(*n*=253) | **G2: Cannabis Use, Not for Mental Health**<br>(*n*=131/94) | **G3: Cannabis Use, for Mental Health**<br>(*n*=253/131) |
| --- | --- | --- | --- |
| **BASELINE Model** |  |  |  |
| Intercept | 1.54*** | 1.18*** | 2.37*** |
| Slope | -0.36** | -0.05 | -0.68* |
| Slope-Intercept<br>Correlation | -0.35** | -0.68*** | -0.27 |
| **FINAL Model** |  |  |  |
| Intercept | 1.56*** | 1.63*** | 2.30*** |
| Slope | -0.43*** | 0.00 | -0.51* |
| Slope-Intercept<br>Correlation | -0.36** | 0.00 | -0.28 |

**Table 6.** COPE Model 2: Does Stress Decrease Among Moms Using Cannabis to Cope with Mental Health Difficulties?

|  | **G1: No Cannabis Use**<br>(*n*=223) | **G2: Cannabis Use, Not for Mental Health**<br>(*n*=78) | **G3: Cannabis Use, for Mental Health**<br>(*n*=116) |
| --- | --- | --- | --- |
| **BASELINE Model** |  |  |  |
| Intercept | 2.5*** | 2.26*** | 3.36*** |
| Slope | -0.34* | -0.04 | -0.37* |
| Slope-Intercept<br>Correlation | -0.41** | -0.50*** | -0.69*** |

| **FINAL Model** |  |  |  |
| --- | --- | --- | --- |
| Intercept | 1.56*** | 2.92*** | 3.34*** |
| Slope | -0.43*** | 0.00 | -0.32** |
| Slope-Intercept<br>Correlation | -0.36** | 0.00 | -0.69*** |

## Discussion

This study characterized trajectories of cannabis use, depressive symptoms, and stress symptoms among individuals at higher likelihood for both cannabis use and mental health symptoms during the prenatal period. Examination of mean symptoms at each trimester revealed that cannabis use, depression, and stress decreased for all moms. We attempted to elucidate drivers of these decreases by analyzing these trajectories within a series of latent growth curve models. Baseline, covariate-adjusted models showed that first trimester PCU was higher in those who were younger and had greater levels of poverty. Not surprisingly, mental health history was associated with higher depressive symptoms in the first trimester and with less change across time. First trimester prenatal stress was associated with younger age and a history of mental health conditions.

Inclusion of mental health motives for use – in other words, mothers who cited using cannabis during pregnancy as a way to cope with depression and anxiety – into the growth models showed that first trimester cannabis use was associated with higher rates of depressive and stress symptoms among those using for mental health reasons. Notably, while there were decreases in depressive and stress symptoms across pregnancy among mothers using cannabis for mental health reasons, the rate of change was equivalent to mothers who were not using cannabis during pregnancy. Meanwhile, mothers using cannabis for alternate reasons had a relatively flat distribution of depressive and stress symptoms in that these mothers did not have high symptoms at baseline nor did they change much by the third trimester.

The parallel process model revealed that both the intercepts and slopes (rate of change) between cannabis use and depressive symptoms were correlated with one another. Despite this correlation, first trimester cannabis use did not affect depression’s negative slope across the prenatal period. Likewise, first trimester depression did not affect the slope for cannabis use across the prenatal period. The covariate-adjusted parallel models were similar to the baseline models, in that age and mental health history were associated with T1 depression, while age and INR were associated with T1 cannabis use. Mental health history was associated with a relatively slower decrease in depressive symptoms over time, and was nominally significantly associated with decreased cannabis use over time. The parallel process model for cannabis use and stress showed that the intercepts, but not slopes, were correlated with one another. As in the depression parallel model, age and mental health history were associated with T1 stress, while age and INR were associated with T1 cannabis use. In contrast to the depression model, mental health history did not affect the slope of stress or cannabis use.

Among the participants in our sample, duration of pregnancy seemed to have a protective effect against stress and depressive symptoms, as well as intensity of cannabis use. Extant literature on the effect of timing on depressive symptoms during pregnancy has been mixed; some studies, like ours, have reported a protective effect of time (Dekel et al., 2019), while others have reported that those who begin pregnancy at a high rate of depression tend to remain high throughout their pregnancy (Castro et al., 2017). Other studies have found that, while rates of depression decrease from the prenatal to the postpartum period, rates tend to be fairly stable over the prenatal timepoints (Heron et al. 2004; Boekhorst et al., 2019).

A core component of the present study was ascertainment of reasons why pregnant individuals chose to continue cannabis use following knowledge of their pregnancy. Indeed, the majority of studies on cannabis use during pregnancy focus disproportionately on its effects on fetal development and birth outcomes. While it is undoubtedly important to understand the effect of maternal cannabis use on infant development, targeted interventions to decrease prenatal cannabis use, without a thorough understanding of the function of this behavior, may not be effective. One recent review identified six studies that examined pregnant individuals’ attitudes toward cannabis use during pregnancy and found that participants primarily cited using to treat nausea (Bayrampour et al., 2019). Further, this study found that individuals also perceived low risk involved with prenatal cannabis use due to a lack of counseling from their obstetric providers.

While some adult cannabis users report using to mitigate the effects of anxiety and depression, multiple studies have in fact found that consistent cannabis use is associated with a wide spectrum of psychiatric conditions in long-term users (Kedzior et al. 2014; Gobbi et al. 2019). This discordance may be due to the short-term relief many experience during use (Black et al. 2019). This may be reflective of the findings in our study; while self-reported stress and anxiety decreased among cannabis users with motivation for mental health reasons, the rate of change was identical to mothers using cannabis for other reasons. Between these findings and the previously cited literature which points to a lack of counseling around cannabis use from obstetric providers, it is important to clarify the lack of evidence for significant and sustained mental health symptom relief as a result of cannabis use to pregnant individuals.

### Strengths / limitations

This study has several strengths. Importantly, this is one of the only samples to date to recruit women who were engaging in cannabis use separately from nicotine use. Thus, we are able to draw conclusions about the effects of cannabis use distinct from any potential confounding effects as the result of co-occurring nicotine use. Additionally, women’s self-reports of cannabis use were verified throughout their pregnancy with the addition of consistent urine drug screen tests.

The study should be interpreted in light of some limitations. Because we were interested in interactions among motives for cannabis use, cannabis use group, and mental health status, we were slightly underpowered.

## Conclusions

We investigated trajectories of depressive symptoms, stress burden, and cannabis use across the prenatal period. While depression, stress, and cannabis use all decreased from the first to the third trimester, cannabis use at baseline was not associated with the rate of change in depressive symptoms. Likewise, depression at baseline was not significantly associated with the rate of change in cannabis use. In contrast, baseline depression was associated with the rate of change in stress and vice versa. Mothers using cannabis for mental health reasons should be informed that cannabis does not appear to have a measurable effect on the rate of change in mental health symptoms during the prenatal period.

## Data Availability

All data produced in the present study are available upon reasonable request to the authors

## References

Agrawal A, Rogers CE, Lessov-Schlaggar CN, Carter EB, Lenze SN, Grucza RA. Alcohol, cigarette, and cannabis use between 2002 and 2016 in pregnant women from a nationally representative sample [published online November 5, 2018]. JAMA Pediatr. doi:10.1001/jamapediatrics.2018.3096

Besse, M., Parikh, K., & Mark, K. (2023). Reported Reasons for Cannabis Use Before and After Pregnancy Recognition. Journal of addiction medicine, 17(5), 563–567.

Black, N., Stockings, E., Campbell, G., Tran, L. T., Zagic, D., Hall, W. D., … & Degenhardt, L. (2019). Cannabinoids for the treatment of mental disorders and symptoms of mental disorders: a systematic review and meta-analysis. The Lancet Psychiatry, 6(12), 995–1010.

Bollen, K. A., & Curran, P. J. (2006). Latent curve models: A structural equation perspective. John Wiley & Sons.

Cohen, S., Kamarck, T., & Mermelstein, R. (1994). Perceived stress scale. Measuring stress: A guide for health and social scientists, 10(2), 1–2.

Cox, J. L., Holden, J. M., & Sagovsky, R. (1987). Detection of postnatal depression: development of the 10-item Edinburgh Postnatal Depression Scale. The British journal of psychiatry, 150(6), 782–786.

Dennis, C. L., Falah-Hassani, K., & Shiri, R. (2017). Prevalence of antenatal and postnatal anxiety: systematic review and meta-analysis. The British Journal of Psychiatry, 210(5), 315–323.

Ding, X. X., Wu, Y. L., Xu, S. J., Zhu, R. P., Jia, X. M., Zhang, S. F., … & Tao, F. B. (2014). Maternal anxiety during pregnancy and adverse birth outcomes: a systematic review and meta-analysis of prospective cohort studies. Journal of affective disorders, 159, 103–110.

El Marroun, H., Tiemeier, H., Jaddoe, V. W., Hofman, A., Mackenbach, J. P., Steegers, E. A., … & Huizink, A. C. (2008). Demographic, emotional and social determinants of cannabis use in early pregnancy: the Generation R study. Drug and alcohol dependence, 98(3), 218–226.

Fedock, G. L., & Alvarez, C. (2018). Differences in screening and treatment for antepartum versus postpartum patients: are providers implementing the guidelines of care for perinatal depression?. Journal of Women’s Health, 27(9), 1104–1113.

Field, T. (2017). Prenatal anxiety effects: a review. Infant Behavior and Development, 49, 120–128.

Gobbi, G., Atkin, T., Zytynski, T., Wang, S., Askari, S., Boruff, J., … & Mayo, N. (2019). Association of cannabis use in adolescence and risk of depression, anxiety, and suicidality in young adulthood: a systematic review and meta-analysis. JAMA psychiatry, 76(4), 426–434.

Hammond, D., Goodman, S., Wadsworth, E., Rynard, V., Boudreau, C., & Hall, W. (2020). Evaluating the impacts of cannabis legalization: the International Cannabis Policy Study. International Journal of Drug Policy, 77, 102698. JAMA. 2018;320(6):545–546. doi:10.1001/jama.2018.8401

Kedzior, K. K., & Laeber, L. T. (2014). A positive association between anxiety disorders and cannabis use or cannabis use disorders in the general population-a meta-analysis of 31 studies. BMC psychiatry, 14, 1–22.

Kiel, L., Hsu, C., Wartko, P. D., Albertson-Junkans, L., Ewing, J., & Lapham, G. T. (2023). Perspectives from women who engaged in prenatal and postpartum cannabis use in a US State with legal non-medical use. Preventive Medicine Reports, 31, 102075.

Ko, J. Y., Farr, S. L., Tong, V. T., Creanga, A. A., & Callaghan, W. M. (2015). Prevalence and patterns of marijuana use among pregnant and nonpregnant women of reproductive age. American journal of obstetrics and gynecology, 213(2), 201–e1.

Lauren M. Jansson, MD1; Chloe J. Jordan, PhD2; Martha L. Velez, MD1 Mateus, V., Cruz, S., Costa, R., Mesquita, A., Christoforou, A., Wilson, C. A., … & Osório, A. (2022). Rates of depressive and anxiety symptoms in the perinatal period during the COVID-19 pandemic: Comparisons between countries and with pre-pandemic data. Journal of affective disorders, 316, 245–253.

Pearson, R. M., Carnegie, R. E., Cree, C., Rollings, C., Rena-Jones, L., Evans, J., … & Lawlor, D. A. (2018). Prevalence of prenatal depression symptoms among 2 generations of pregnant mothers: the Avon longitudinal study of parents and children. JAMA network open, 1(3), e180725–e180725.

Perinatal Marijuana Use and the Developing Child Regalado, D., Connolly, M. E., Krutsch, K., Stark, A., Kendall-Tackett, K., & Garner, C. D. (2023). Psychiatric medication use among pregnant and breastfeeding mothers who used cannabis for mental health concerns: A cross-sectional survey study. Women’s Health, 19, 17455057231199391.

Rubertsson, C., Hellström, J., Cross, M., & Sydsjö, G. (2014). Anxiety in early pregnancy: prevalence and contributing factors. Archives of women’s mental health, 17, 221–228.

Singh, S., Filion, K. B., Abenhaim, H. A., & Eisenberg, M. J. (2020). Prevalence and outcomes of prenatal recreational cannabis use in high-income countries: a scoping review. BJOG: An International Journal of Obstetrics & Gynaecology, 127(1), 8–16.

Skelton, K. R., Hecht, A. A., & Benjamin-Neelon, S. E. (2021). Association of recreational cannabis legalization with maternal cannabis use in the preconception, prenatal, and postpartum periods. JAMA network open, 4(2), e210138–e210138.

Vanstone, M., Taneja, S., Popoola, A., Panday, J., Greyson, D., Lennox, R., & McDonald, S. D. (2021). Reasons for cannabis use during pregnancy and lactation: a qualitative study. Cmaj, 193(50), E1906–E1914.

Volkow ND, Compton WM, Wargo EM. The risks of marijuana use during pregnancy. JAMA. 2017;317(2):129–130. doi:10.1001/jama.2016.18612

Woody, C. A., Ferrari, A. J., Siskind, D. J., Whiteford, H. A., & Harris, M. G. (2017). A systematic review and meta-regression of the prevalence and incidence of perinatal depression. Journal of affective disorders, 219, 86–92.

